# CLINICOPATHOLOGIC FEATURES OF URINARY BLADDER CANCER AT A TERTIARY HOSPITAL IN WESTERN KENYA

**DOI:** 10.1101/2024.05.10.24307172

**Authors:** Charles Sore Oduor, Edward Mugalo, Geoffrey Kirongo

## Abstract

**Background:** Urinary bladder cancer is the ninth leading cause of morbidity and mortality globally, with a prevalence of 3% of all cancer diagnoses. Its local prevalence in Kenya is 1.89 per 100, 000 persons with previously documented studies describing it as a disease of the elderly. Local anecdotal data indicate that younger patients have begun presenting with bladder cancer.

**Objectives:** To establish the clinicopathological features of urinary bladder cancer.

**Methods:** A prospective descriptive hospital-based study among participants with bladder cancer at MTRH Urology department, sampled using a census sampling technique. Socio-demographic and clinical characteristics as well as predisposing factors were obtained through both interviews and a review of medical records. Disease staging was based on radiological imaging findings and histopathology reports. Statistical tests of association between socio-demographic characteristics, predisposing factors as well as the histological type, and Tumor, Node, Metastasis (TNM) stage of urinary bladder cancer were conducted using Pearson chi-square test with a critical value of ≤0.05.

**Results:** Forty-five (45) adults aged between 21 to 85 years with a mean age of 61.84 (±14.46) years and diagnosed with urinary bladder cancer were enrolled. Majority were male (68.9%; n=31) commonly presenting with painless hematuria, exposure to agrochemicals (60%; n=27), history of cigarette smoking (31.1%; n=14) with an average pack year of 9.43 (±6.198). The most common (71.1%; n=32) clinical stage was T_1_ while 6.7% (n=3) had metastatic disease. More than half (55.6%) had stage I according to the TNM system while 91.1% had low-grade tumors. Transitional cell carcinoma (51.1%) was the most common histological type, followed by adenocarcinoma (29%), and squamous cell carcinoma (20%).

**Conclusions:** Patients diagnosed with bladder cancer in this study were mainly males with a mean age of 61.8 years. Majority of the participants had a history of smoking cigarettes or exposure to agrochemicals, and all presented with painless hematuria. Most patients had low-grade tumors diagnosed early.

## Introduction

Bladder cancers are among the most common malignant neoplasms that involve the urinary tract system^1^. Urinary bladder cancer has been ranked the ninth among most prevalent malignant diseases and it is ranked thirteenth among cancer-related deaths globally ^2^. Its prevalence has been estimated at 3% of all cancer diagnoses globally ^3^ with a prevalence of 1.2% ^4^ in North America. Its incidence in the Eastern Africa region is estimated at 2.8% with the male and female incidence at 3.2% and 2.4% respectively^1^. Despite this high disease burden and mortality risk, urinary bladder cancer receives limited attention from both healthcare policymakers and healthcare practitioners in comparison to prostate cancer among men and breast and cervical cancer among women^5^.

Bladder cancer is the sixth most common urologic malignancy globally and ninth commonest in Kenya^5,6^. It is associated with elevated morbidity and increased rates of mortality rates if not well treated ^7,8^. Majority of the patients diagnosed with bladder cancer are older males, with a history of smoking or occupational exposure to carcinogens, chronic cystitis particularly with schistosomiasis, first-degree relatives (parent, sibling, or child) with a family history of urinary bladder cancer or those who have undergone previous cancer treatment ^4,9,10^. These at-risk individuals are often bread winners whose quality of life and socioeconomic activities have been interrupted by the disease ^11^. This not only affects them at the individual level but the community as a whole. The hospital records at MTRH indicate that patients younger than 30 years with no associated familial history and identifiable exposure to carcinogens have been diagnosed with various types of urinary bladder cancer. Additionally, there are multiple histological bladder types of urinary bladder cancer with varying prevalence and prognosis depending on the predisposing factors. This necessitates a better understanding of clinicopathological features, mode and stage at presentation as well as treatment approaches. Although there is the possibility of clinical management of the disease, treatment resistance risks abound enabling a combined approach that integrates surgery. The preferred mode of management of muscle-invasive bladder cancer is surgical, however, empirical data indicate several challenges that are not limited to high rates of recurrence and surgical complications that directly impact the patient’s quality of life. Multiple published studies have reported late presentation among patients with advanced bladder cancer in countries with developed economies^12,13^. With the foregoing, there is a need for local contextualized data that describes the patient characteristics of those diagnosed with urinary bladder cancer, their predisposing factors, clinical as well as histological variants, and stage at presentation. This is because clinicopathological characteristics have a direct influence on surgical management approaches and treatment outcomes. It is because of this that this study was conducted at Moi Teaching and Referral Hospital (MTRH) urology department which receives a heterogeneous and diverse group of patients from the greater western, northern, and Rift Valley regions of Kenya. This study therefore aims to describe clinicopathological features of urinary bladder cancers at Moi Teaching and Referral Hospital (MTRH).

Urinary bladder cancers are among the major oncologic conditions affecting adults of advanced age with a higher incidence among the males. Despite this knowledge, the clinicopathological features such as patient characteristics, their clinical and histologic stage of presentations have not been well documented. This could be attributed to the fact that most of the studies conducted locally tend to address the prevalence and incidence of urinary bladder cancer. Additionally, although the burden of urinary bladder cancers is known, there are limited local studies addressing specific patient characteristics and their associated factors among those presenting with the disease that have been conducted at a major tertiary facility in Kenya. This study was therefore conducted at Moi Teaching and Referral Hospital (MTRH) which is in Eldoret town and is a major tertiary hospital in Kenya that serves the greater Western region. It addressed the patient sociodemographic and clinical characteristics, associated factors, histopathological variants and stage at presentation. These were done to address the emerging incidence of urinary bladder cancer among younger adults aged below 30 years ^14^. Furthermore, anecdotal data indicate that there is a high prevalence of squamous cell carcinoma in countries such as Kenya which are located along the Lake Victoria Basin and River Nile Banks ^15^. The distribution was associated with increased incidence of schistosomal infection that results in chronic irritation and squamous cell transformation.

Furthermore, knowledge on the commonly presenting clinical stage is needed in both treatment planning and disease control. This knowledge will inform both intervention and control programs of the disease. The findings of this study will inform both clinical care practitioners and policy makers on identifying patient factors associated with the pattern of presentation to inform treatment planning and prevention measures for urinary bladder cancers. The findings of this study will also contribute to both the local and global body of knowledge on urinary bladder cancer.

This study aimed to establish the clinicopathological features of urinary bladder cancer at Moi Teaching and Referral Hospital, Eldoret-Kenya. Specifically, it described their socio-demographic characteristics and clinical features, assessed histological types and clinical stage and analyzed associated factors.

## Methodology

This was a prospective descriptive hospital-based study among patients diagnosed with bladder cancer reviewed at MTRH. The study enrolled 45 adult patients admitted to the surgical urology wards or attending the urology outpatient clinics with urinary bladder cancer identified histologically and staged by radiological imaging. A written informed consent was obtained prior to commencing the study, in a private room by trained research assistants who explained the scope and nature of the study. Participants’ socio-demographic and clinical characteristics data were collected using an interviewer administered structured questionnaire and review of existing medical records (medical charts, histopathology and CT-scan reports). After reviewing data for completes and consistency, continuous variables such as age, the findings were summarized as mean (with corresponding standard deviations) as well as median (with corresponding interquartile range). Categorical data such as predisposing factors to urinary bladder cancer, histological types, clinical stage at presentation as well as surgical and clinical interventions offered were descriptively summarized as frequencies (with corresponding proportions). Tests of association using Pearson Chi-Square tests as well as multivariate logistic regression were used to test whether there was any statistically significant association between socio-demographic characteristics, predisposing factors as well as histological type or tumor, node and metastasis (TNM) staging of urinary bladder cancer diagnosed at MTRH. A critical value of p≤0.05 was considered statistically significant. Relative Risks were computed at 95% confidence interval using Statistical Package for Social Sciences (SPSS) version 26. Ethical clearance was obtained from the Institutional Research and Ethics Committee (IREC) of Moi University School of Medicine and MTRH (# 0004130). A research license was obtained from the National Commission for Science, Technology and Innovation (NACOSTI /P /22 /18168).

## Results

This study enrolled 45 individuals diagnosed with urinary bladder cancer in MTRH who were aged between 21 to 85 years (with a mean age of 61.84±14.46 years). More than two thirds (68.9%; n=31) of the participants were male and 14(31.1%) were female. More than half (55.6%; n=25) had attained primary education, followed by secondary (33.3%; n=15) and tertiary (11.1%; n=5). When participants’ occupation was assessed, 26 (57.8%) reported that they were unemployed as at the time of data collection with the rest (42.2%; n=19) being employed. About one-third (31.1%; n=14) smoking history with 31.1% (n=14) admitting having smoked with an average of 9.43±6.198 pack years while exposure to agricultural chemicals among 27 (60.0%). All the participants had hematuria, while other commonly noted clinical features were lower abdominal pain (93.3%; n=42), irritative (95.6%; n=43) and obstructive voiding symptoms (91.1%; n=41). The constitutional symptoms noted were weight loss (93.3%, n=42), fatigue (95.6%; n=43), anorexia (91.1%; n=41), night sweats (86.7%; n=39). Lastly, 25 (55.6%) participants presented with a pelvic mass.

The most common histological type noted was transitional cell carcinoma (51.1%), followed by adenocarcinoma (28.9%) and squamous cell carcinoma (20.0%). Of all the participants enrolled 41 (91.1%) had low grade and 4 (8.9%) had high grade tumors (Figure 4.1).

**FIGURE 4.1:**
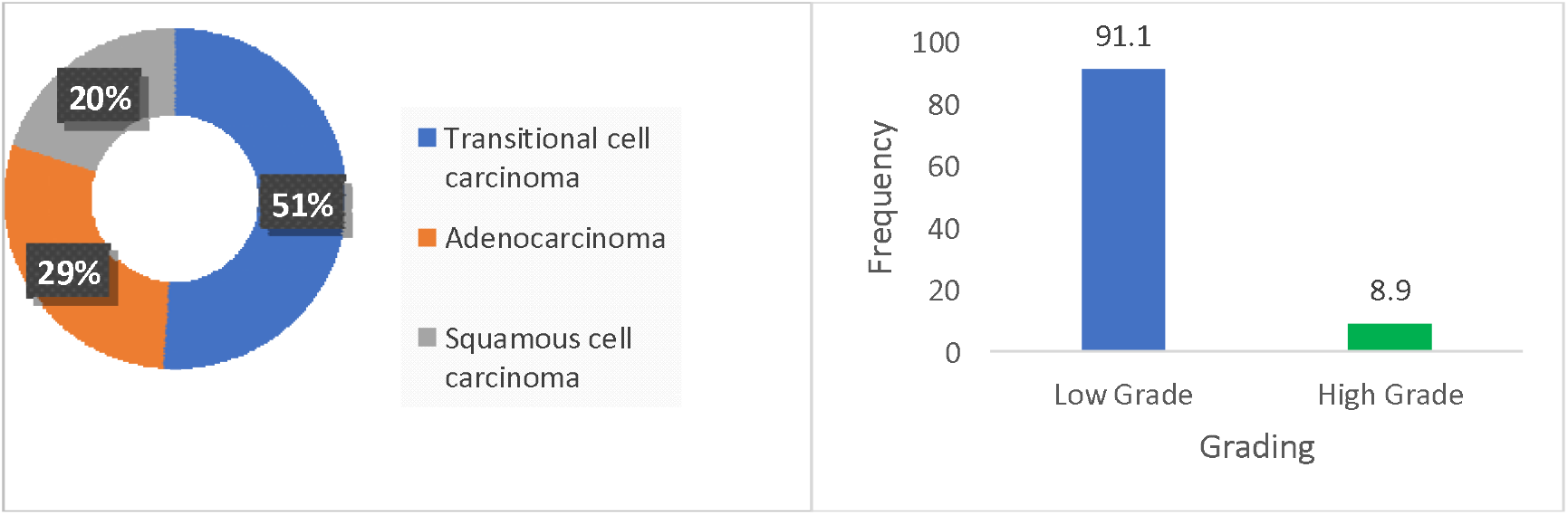
HISTOLOGICAL TYPES AND GRADE OF URINARY BLADDER CANCER All the study participants had multiple lesions which appeared intraoperatively as fragments (20%), papillary fronds (57.8%), patches (17.8%), polypoid masses (2.2%) and spiral necrotic (2.2%). When the participants were staged clinically, the most common stage was T_1_ (71.1%), T_2a_ (17.8%), T_3_ (8.9%) and T_4a_ (2.2%). 7 (15.6%) had extravesical extensions, 5 (11.1%) had nodal involvement of the disease, 3 (6.7%) metastatic disease. As per the TNM staging criteria, the most prevalent was Stage I (55.6%), followed by Stage II (31.1%) and Stage IIIa at 6.7%(Table 4.2).

**Table 4.1:** Participants’ sociodemographic characteristics.

| Sociodemographic Characteristics |  | n (%)/ Mean (SD) |
| --- | --- | --- |
| Age (years) | Less than 30 | 2 (4.4) |
|  | 30-39 | 1 (2.2) |
|  | 40-49 | 7 (15.6) |
|  | 50-59 | 5 (11.1) |
|  | 60-69 | 16 (35.6) |
|  | Above 70 | 14 (31.1) |
| Sex | Male | 31 (68.9) |
|  | Female | 14 (31.1) |
| Level of Education | Primary | 25 (55.6) |
|  | Secondary | 15 (33.3) |
|  | Tertiary | 5 (11.1) |
| Employment status | Employed | 19 (42.2) |
|  | Unemployed | 26 (57.8) |
| Predisposing Factors | Smoking History ($n=14$; 31.1%) | Pack years ($9.43 \pm 6.198$) |
|  | Exposure to Agrochemicals | 27 (60.0) |
| Hematuria | Yes | 45 (100.0) |
| Abdominal Pain | Yes | 42 (93.3) |
| Irritative voiding Symptoms | Dysuria | 42 (93.3) |
|  | Increased frequency of urination | 41 (91.1) |
|  | Urge incontinence. | 39 (86.7) |
|  | Nocturia | 41 (91.1) |
| Obstructive Voiding Symptoms | Straining | 37 (82.2) |
|  | Intermittent stream | 39 (86.7) |
|  | Poor stream of urine | 35 (77.8) |
|  | Feeling of incomplete voiding | 41 (91.1) |
| Constitutional Symptoms | Weight Loss | 42 (93.3) |
|  | Fatigue | 43 (95.6) |
|  | Anorexia | 41 (91.1) |
|  | Night sweats | 39 (86.7) |
|  | Pelvic mass | 25 (55.6) |

**Table 4.2:** Factors associated with histopathological variants of urinary bladder cancer.

| Variable | Sub-category | n (%) |
| --- | --- | --- |
| Multiple Lesions |  | 45 (100) |
| Intraoperative Findings (Appearance) | Fragments | 9 (20.0) |
|  | papillary fronds | 26 (57.8) |
|  | Patches | 8 (17.8) |
|  | Polypoid masses | 1 (2.2) |
|  | spiral necrotic | 1 (2.2) |
| Clinical Staging | T<sub>1</sub> | 32 (71.1) |
|  | T<sub>2a</sub> | 8 (17.8) |
|  | T<sub>3</sub> | 4 (8.9) |
|  | T<sub>4a</sub> | 1 (2.2) |
| Imaging Findings | extravesical extensions | 7 (15.6) |
|  | Nodal involvement | 5 (11.1) |
|  | metastases | 3 (6.7) |
| TNM Staging Criteria | Stage 0<sub>is</sub> | 1 (2.2) |
|  | Stage I | 25 (55.6) |
|  | Stage II | 14 (31.1) |
|  | Stage III | 3 (6.7) |
|  | Stage IV<sub>a</sub> | 1 (2.2) |
|  | Stage IV<sub>b</sub> | 1 (2.2) |

After conducting a logistic regression analysis on the relationship between participant socio-demographic characteristic (independent variable) and urinary bladder cancer variants (dependent variable) while controlling for associated factors (mediating variable); this study reports the following statistically significant associations: Male participants who had a history of exposure to agrochemicals were significantly more likely to present with low grade tumors (p=0.013; AOR=2.276, 95% CI: 1.366-9.935) and TNM clinical stage I (p=0.021; AOR=1.737; 95% CI: 1.407-7.658). Furthermore, male participants had a marginally higher risk (p=0.047; RR=1.441; 95% CI: 1.129-5.870) of squamous cell carcinoma of the bladder compared to their female counterparts. Lastly, participants older than 60 years of age (irrespective of their sex) who were exposed to agrochemicals were significantly more likely (p=0.029; RR=2.100, 95% CI: 1.436-9.000) to be diagnosed with adenocarcinoma of the bladder compared to their younger counterparts (Table 4.3).

**Table 4.3:** Factors associated with histopathological variants of urinary bladder cancer.

| Sociodemographic Characteristic | Risk Factor | Histopathological variants of Urinary Bladder Cancer | p-value | AOR (95% CI) |
| --- | --- | --- | --- | --- |
| Sex (Male) | Exposure to Agrochemicals | Low grade Tumor | 0.013 | 2.276 (1.366, 9.935) |
|  |  | TNM Stage I | 0.021 | 1.737 (1.407, 7.658) |
| Age (> 60 years) | Exposure to Agrochemicals | Adenocarcinoma | 0.029 | 2.100 (1.436, 9.000) |

## Discussion

This study enrolled a predominantly higher proportion of male participants (68.9%) compared to females (31.1%). This could be attributed to the fact that impaired emptying of the bladder in male gender that increases with age compared to females. Similar finding was reported in Turkey^16^ where 62% of the participants were male with 38% being female. Similar trends were reported in other studies conducted in Germany, China, Netherlands and the United States of Africa^17–21^. In Germany^17^, 85.7% of those enrolled were male with the rest being female, while in China^18^ 70.3% were male with 29.7% females. In Amsterdam – Netherlands ^19^, 74% were male and 26% female. Urinary bladder cancer was also noted in Nebraska-Omaha, United States of America^22^, to be more predominant among male participants compared to their female counterparts. Additionally, this male predominance of urinary bladder cancer could be attributed to the unhealthy lifestyle of men compared to women living in western Kenya. These include smoking and exposure to agrochemicals which increases the risk of developing urinary bladder cancer among male participants. When the participants’ age was assessed, the mean age of this study’s participants was 61.84 (±14.46) years. A finding that was close to that reported in many studies under comparison as urinary bladder cancers tend to manifest among adult individuals of advanced age ^16,17,20,23^. This advancement of age could also be attributed to the latent effects of the risk factors for bladder cancer such as environmental or dietary exposures, when the symptoms are noticed^19^. The mean age at diagnosis of other studies reviewed was 60.8 ± 14.04 years in Xiamen -China^22^ and 63.2 years in Turkey^16^.

The common clinical features assessed in this study were painless gross hematuria, lower abdominal pain, irritating and obstructive voiding symptoms as well as non-specific constitutional symptoms. In this study, all participants had painless gross hematuria which is a major hallmark of urinary bladder cancer. This finding is comparable to that reported at the University of Nairobi in Kenya^24^ where nearly all (98%) of the participants had painless gross hematuria. A similar finding was reported at the University of Lucknow in India^25^ where 93.3% of the patients presented with painless gross hematuria. Lower hematuria proportions of this were reported in China at 78.3%^20^, 74.5% in Iran^26^, and 72.6% in Pakistan^27^. Nearly all (93.3%) of the patients presented with lower abdominal pain, a finding that matched a previous study conducted in Kenya at 71%^24^. This high proportion could be attributed to the phenomenon that most patients will come to the hospital complaining of a painful lower abdomen which on further investigations could be found to be a urinary bladder tumor. Additionally, the commonly assessed irritative voiding symptoms in this study were dysuria, increased frequency of urination and urge incontinence. This study reports that 93.3% of its participants had dysuria. This is a condition that is associated with painful micturition (passage of urine). Because of the nature and location of urinary bladder tumors, the bladder is squeezed, and this increases peripheral resistance during the passage of urine and the associated pain. The findings of this study on dysuria are higher than all the studies under comparison at 66.7% in India^28^, Kenya^24^ at 31% and 5.5% in Iran^26^. Additionally, 91.1% of participants in this study complained about the increased frequency of urination, a proportion higher than all studies under comparison from Iran^26^, India^28^, and Pakistan^27^ which reported proportions of 41.5%, 36.7%, and 22.1% respectively. Lastly, more than three-quarters (86.7%) of the current study’s participants reported urge incontinence a proportion which was about three times higher than that reported in India at 33.3%^28^. This high proportion of patients with urge incontinence could be due to associated urinary tract infections.

Multiple patient factors could increase the risk of an individual developing urinary bladder cancer. In this study, about one-third (31.1%) of the study participants were found to have a history of smoking with an average of 9.43 (±6.198) pack years. In other comparative studies^29–31^, half of the participants in the United States of America and Korea reported a smoking history. A higher proportion (73.4%) of reported smoking history was noted in India^28^ where about three-quarters of the study participants reported a history of smoking. This proportionate difference could be attributed to sociocultural and demographic differences, as a higher proportion of smoking has been reported in countries with higher mean income located in the continents of America, Europe and Asia. Furthermore, this study notes exposure to agrochemicals as a major risk factor for urinary bladder cancer. Because this study was conducted in Eldoret town, which is predominantly an agricultural area as it is the breadbasket of Kenya, it could explain why nearly two-thirds (60%) of the participants diagnosed with urinary bladder cancer had an occupational exposure to agrochemicals. Previous studies ^32,33^ have argued that exposure to agricultural chemicals causes cellular transformation which may interfere with the cell cycle especially the apoptotic process. This is because these agrochemicals are considered to be carcinogens. Just like agrochemicals, 2.2% of those enrolled in this study were painters and would be constantly exposed to oil paints in the course of their seeking a livelihood. The active ingredients of paints as well as their excipients have been noted to be carcinogens, and when inhaled over a prolonged period, there is a great risk of cancer manifestation^34^.

When the study assessed the most common histological types of urinary bladder cancer identified in this study were transitional cell carcinoma (51.1%), Adenocarcinoma (28.9%) and squamous cell carcinoma (20.0%). The findings on transitional cell carcinoma were close to that reported in Kenya^24^ at 67%. This could be attributed to the fact that both studies were conducted in the same country where clinical findings follow a sociodemographic pattern. However, the slight proportionate difference could be attributed to temporal differences. This study was conducted in the year 2022 while Waihenya and colleagues published their findings in the year 2004. Higher proportions of transitional cell carcinoma were reported in two studies conducted in India at 90.5%^27^, 97.6%^35^and Libya^36^ at 87.4%. Transitional cell carcinoma of the bladder has been associated mainly with cigarette smoking and myriad of occupational exposures. The trend may worsen in the near future in developing countries if measures to control cigarette smoking (or other nicotine derivatives) are not put in place. The proportion of adenocarcinoma reported in this study is more than three folds higher than that previously reported in Kenya at 8%^24^. Whereas the current study adopted a prospective design, that was conducted in Nairobi adopted a ten-year retrospective design which is often fraught with incomplete data that may cause proportionate variations on the factors of interest. Squamous cell carcinoma proportion in this study could be attributed to chronic urinary tract infections from either bacteria or *Schistosoma haematobium* parasite which are prevalent in Western Kenya which is within the Lake Victoria basin^10,37^.

This study reported that nine in ten (91.1%) of the participants had low grade tumors with 8.9% presenting with high grade tumors. This could be attributed to early patient presentation at first encounter as was also the case in Libya^36^ where 83.4% had a low tumor grade. However, contrasting histological grade findings could be due to the varied patient characteristics and the reported risk factors contributing to the disease. Because of the slow manifestations of many oncologic conditions including urinary bladder tumors, there is a great likelihood of late diagnosis in many countries, especially those with developing economies. Low-grade tumors are well differentiated with a good prognostic feature and reduced risk of recurrence.

The clinical stages of interest were classified as either non-muscle or muscle-invasive tumors. Specifically, 32 (71.1%) of the participants had non-muscle invasive tumors while muscle invasive tumors were presented as Stage T2a (17.8%), Stage T3 (8.9%) and Stage T4a (2.2%). The findings of the current study match those presented in a histopathological study conducted in Nepal^35^ where 64.18% of those enrolled had non-muscle invasive clinical stage of urinary bladder tumor identified. This can be described as a sign of early diagnosis with good prognosis if prompt treatment is offered to patients. When the TNM staging criteria were adopted in this study, the most prevalent stage was Stage I at 55.6%, followed by Stage II (31.1%), Stage IIIa (6.7%); with low proportions of Stage 0is, IVa and IVb at 2.2% each. In Egypt^15^, 6.7% of the study participants were diagnosed with Stage 0is, 52.4% had Stage I, 30.2% with Stage II while 7.4% and 3.3% had Stages III and IV respectively Metastatic tumors often spread to distant sites and may lead to secondary tumors, further complicating patient outcomes.

## Conclusion

This study found that the majority of patients diagnosed with bladder cancer at Moi Teaching and Referral Hospital were males aged averagely of 61.8 years. Most of the participants were farmers who interacted with agrochemicals or had a history of smoking cigarettes. The common clinical features of urinary bladder cancer reported in this study were classical symptoms of early disease with most participants presenting with low-grade transitional cell carcinoma followed by adenocarcinoma and squamous cell carcinoma. Most of the patients presented early at diagnosis of the disease, with low proportions of non-muscle invasive tumors. Lastly, male participants exposed to agrochemicals were significantly more likely to present with low-grade tumors and TNM clinical stage I.

Because this study identified that occupational exposure to carcinogens and being of advanced age (older than 60 years) were significantly associated with urinary bladder cancer, there is need to implement early detection strategies for adult males of advanced age. Furthermore, patients presenting with classical symptoms of early disease should be further screened for urinary bladder cancer. From this study, it has been noted that some populations are at higher risk of developing bladder cancer; therefore, there is need for screening to inform the right interventions promptly. Health promotion professionals should also be supported in empowering community members to seek early medical attention for urinary bladder cancer as well as other oncologic conditions, as this study noted that the majority of those enrolled presented early. Early presentation not only reduces the overall cost of care to the affected patients but also improves their overall quality of life and well-being because of better prognosis associated with it.

## Data Availability

All data produced in the present study are available upon reasonable request to the authors

## Acknowledgement

The authors would like to thank the patients who participated in this study as well as the management of Moi Teaching and Referral Hospital.

## Author Contribution

*Conceptualization:* CSO, EM, GK; *Formal Analysis*: CSO, *Resources EM, GK*; *Data Curation:* CSO, EM, GK; *Writing – Original Draft:* CSO.; *Writing – Review & Editing:* CSO, EM, GK; *Supervision:* EM, GK.

